# Can a second booster dose be delayed in patients who have had COVID-19?

**DOI:** 10.1101/2021.04.09.21255200

**Authors:** Jorg Taubel, Christopher S Spencer, Anne Freier, Dorothée Camilleri, Ibon Garitaonandia, Ulrike Lorch

## Abstract

Vaccination forms a key part of public health strategies to control the spread of SARS-CoV-2 globally. In the UK, two vaccines (BNT162b2-mRNA produced by Pfizer, and ChAdOx-1-S produced by Oxford-AstraZeneca) have been licensed to date, and their administration is prioritised according to individual risk. This study forms part of a longitudinal assessment of participants ‘SARS-CoV-2-specific antibody levels before and after vaccination. Our results confirm that there is little quantitative difference in the antibody titres achieved by the two vaccines. Our results also suggest that individuals who have previously been infected with SARS-CoV-2 achieve markedly higher antibody titres than those who are immunologically naïve. This finding is useful to inform vaccine prioritisation strategies in the future: individuals with no history of SARS-CoV-2 infection should be prioritised for a second vaccine inoculation.

## Introduction

SARS-CoV-2 emerged in late 2019 as the causative agent for COVID-19 and over a year later, it remains an ongoing pandemic. Though infection is often asymptomatic, the virus has been shown to cause a range of clinical outcomes including mild infection, severe respiratory distress and death^1^. Vaccination programs have long been held to be the path out of the SARS-CoV-2 pandemic, and in late 2020, the first vaccine for SARS-CoV-2 was licensed in the UK. To date a total of three vaccines have been licensed in the UK, two of which (BNT162b2-mRNA produced by Pfizer-BioNTech and ChAdOx-1-S produced by Oxford-Astra-Zeneca) were made widely available^2^. Both vaccines use a two-dose, prime-boost strategy^3,4^ with vaccine administration focusing on the first of the two inoculations, prioritising individuals according to their risk of severe infection^2^. For each of the two vaccines, the first dose is correlated with lower viral titres and a reduction in symptom severity. The second dose is associated with further reductions in symptom severity and increased viral neutralisation. Previously, it has been shown that upon inoculation with the BNT162b2-mRNA vaccine, individuals with prior SARS-CoV-2 infection produced an order of magnitude more SARS-CoV-2-specific antibodies than those who were uninfected^5^.

This prompted us to examine whether individuals who had previously been infected with SARS-CoV-2 could be deprioritised for receiving a second vaccine inoculation, and whether the answer to this question is dependent on the specific vaccine provided. Therefore, we are conducting an ongoing quantification of the levels of antibodies among our longitudinal cohort of staff at our clinical unit. The results presented herein are made up of eight weeks of post-vaccination data.

## Methods

### Participants and testing methodologies

Sixty-four individuals aged 22 to 63 years received at least one vaccine dose. Twenty-four individuals received the BNT162b2-mRNA vaccine and 40 received the ChAdOx-1-S vaccine. Nineteen vaccinated individuals had previous SARS-CoV-2 infection; (nine received BNT162b2-mRNA vaccine, 10 received ChAdOx-1-S). Those previously infected were vaccinated within an average 6.25 months after infection (range: 2 to 11 months). Cohort demographics are shown in **Table 1**.

**Table 1.** Demographic data of trial participants.

|  | Previous SARS-CoV-2 infection<br>n = 19 | No SARS-CoV-2 infection history<br>n = 45 |
| --- | --- | --- |
| **Vaccine administered, n (%)** |  |  |
| BNT162b2-mRNA | 9 | 15 |
| ChAdOx-1-S | 10 | 30 |
| **Sex, n (%)** |  |  |
| Male | 10 | 16 |
| Female | 9 | 29 |
| **Race, n (%)** |  |  |
| Black African | 1 (5.3) | 1 (2.2) |
| Black Caribbean | 0 (0) | 3 (6.7) |
| Caucasian | 12 (63.2) | 26 (57.8) |
| Chinese | 0 (0) | 1 (2.2) |
| Indian | 0 (0) | 3 (6.7) |
| Japanese | 1 (5.3) | 5 (11.1) |
| Mixed Race | 3 (15.8) | 1 (2.2) |
| Other Asian | 2 (10.5) | 4 (8.9) |
| Taiwanese | 0 (0) | 1 (2.2) |
| **Age (years)** |  |  |
| Mean (SD) | 36.6 (11.5) | 35.6 (11.2) |
| Range | 24 - 63 | 22 - 62 |

### Antibody Quantification

To quantify antibody titres, up to 30 ml of blood was taken from volunteers via venepuncture. Antibody quantification was performed using the anti-S; Elecsys anti-SARS-CoV-2 spike ECLIA Anti-SARS-CoV-2 (Roche Diagnostics, UK) test^6^.

### Statistical Analysis

The data was analysed using summary statistics and overlaid individual plots to determine the differences in antibody responses between individuals who had previously been infected with SARS-CoV-2 and those with no prior infection, and the type of vaccine they received. Due to the nature of the data, the antibody values are log-transformed and the geometric means are calculated. Visual data presentation was done using logscale.

### Ethical Statement

This brief report communicates results taken from a study reviewed and given favourable opinion by the NRES Committee (West Midlands - Edgbaston) (IRAS ID: 281788).

## Results

Irrespective of which vaccine was used, those previously infected produced more antibodies overall and produced antibodies more rapidly. Previously uninfected individuals ‘geometric mean antibody titres were reached 25 days post-vaccination, compared with 16 days for those with prior infection. Four weeks post-vaccination, those with previous infection had antibody titres 59-fold higher than those with no infection history. Comparing the two vaccines, among individuals with no previous infection, the geometric mean titres were almost indistinguishable. For both vaccines, the geometric mean pre-vaccination titre for previously infected individuals was higher than the geometric peak titre for naïve individuals (**Figure 1**).

**Figure 1.**
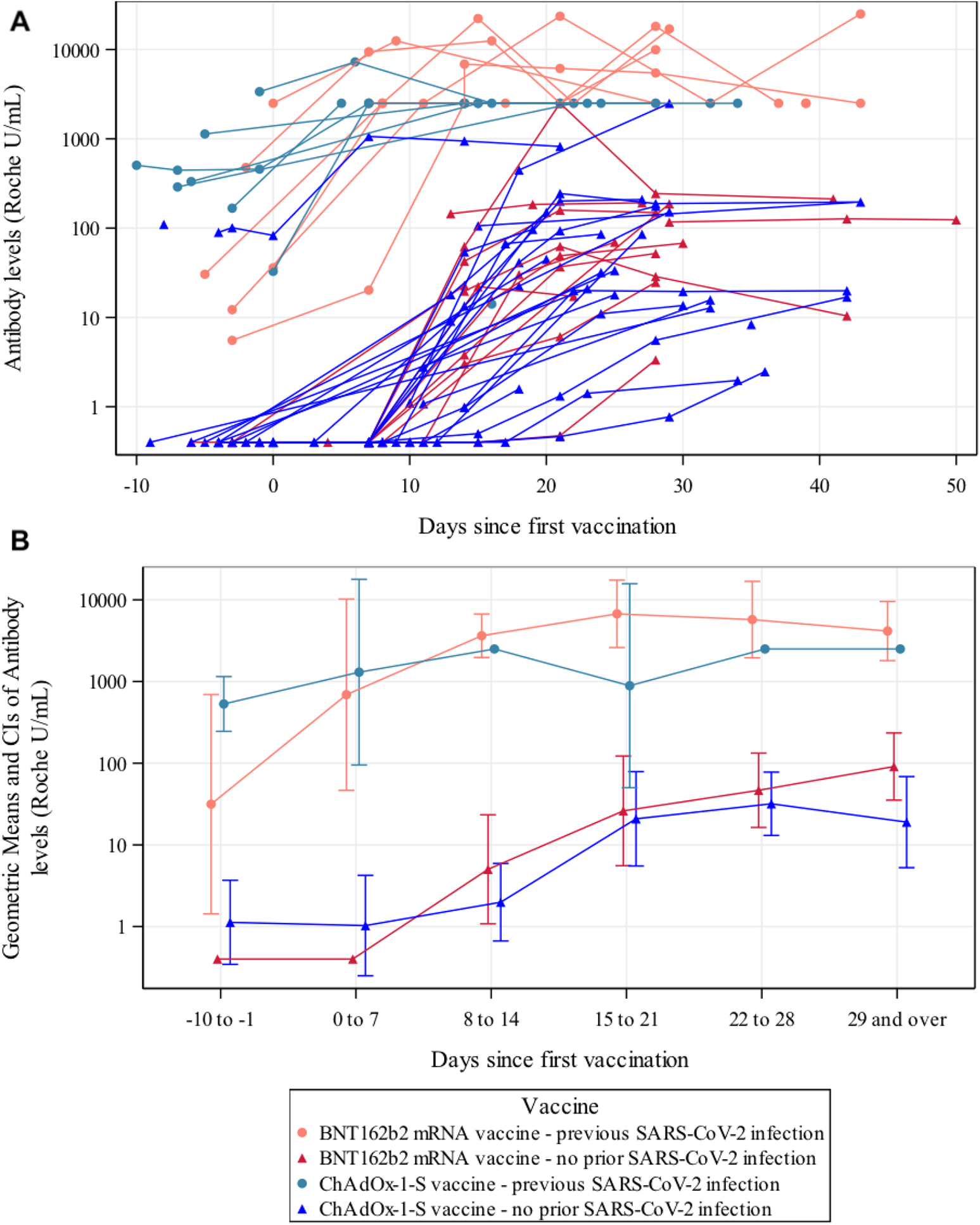
**A)** Serological response to one dose of the BNT162b2 mRNA COVID-19 vaccine (Pfizer) or ChAdOx-1-S vaccine (AstraZeneca) in individuals with and without previous SARS-CoV-2 infection. Individuals with previous infections produce an antibody response faster on average, and with a higher overall titre. Both vaccines give a strong antibody response in infected and uninfected individuals. Among uninfected individuals, there is little difference between the two vaccines. **B)** Geometric mean serological response to one dose of the BNT162b2 mRNA COVID-19 vaccine (Pfizer) or ChAdOx-1-S vaccine (AstraZeneca) in individuals with and without previous SARS-CoV-2 infection.

We found that pain at injection site, chills, headache, and fever were the most common side effects reported. Those receiving ChAdOx-1 appeared more likely to suffer any specific side effect than those receiving the BNT162b2 mRNA vaccine. However, further analysis is required to understand the general applicability of these findings.

An interesting observation is that one individual with no known history of SARS-CoV-2 infection receiving the ChAdOx-1-S vaccine appeared to have antibody levels more consistent with a prior infection. The fact that this person had SARS-CoV-2 antibodies pre-vaccination indicates that they were infected but remained unaware thereof, as there would be no other way for them to have antibodies before vaccination^7^. Our findings suggest that previous infection leads to a higher antibody titre post vaccination, even in previously infected individuals with asymptomatic COVID-19.

## Discussion

Our results showed that previously infected individuals produced a stronger antibody response overall and produced this response more rapidly. In light of these findings, we suggest that a second vaccination for previously infected individuals may not be necessary in the short-term. It may be recommendable to prioritise individuals with no prior history of SARS-CoV-2 infection for a second vaccination, ahead of previously infected individuals who show a stronger response to their first inoculation. Conducting serology on people previously infected to confirm their high antibody titres will allow for a higher number of naïve people to be vaccinated in the short-term.

One limitation of our study was that numerous individuals ‘titres were outside the range of quantification for the ECLIA test. We are therefore unsure of the precise antibody levels. However, sensitivity analysis showed that geometric means allowed us to successfully compare these antibody levels. No individual without prior infection had a titre high enough to exceed the upper bounds of our test, further illustrating that prior SARS-CoV-2 infection yields a much higher antibody titre.

Though this is interim data, over the coming months, we will be able to achieve a more rounded picture of our cohort, who are due to receive their second injections in the coming weeks. Further work is required to examine whether the longevity of the immune response is different based on SARS-CoV-2 infection history. Importantly, whilst we did see some small differences in antibody levels between the two vaccines, we do not believe there to be any appreciable difference in the protection afforded by these two vaccines, in line with previous findings^8^.

## Data Availability

Data are available upon reasonable request.

## Notes

### Competing Interest Statement

The authors have declared no competing interest.

### Clinical Trial

IRAS ID: 281788
NCT04404062

### Funding Statement

This work was self-funded by Richmond Research Institute

### Author Declarations

NRES Committee (West Midlands - Edgbaston)

### Summary of Updates

NCT number included

